# *CARS*2 Hypermethylation Is a Risk Factor for Stage B Heart Failure and Hospitalizations in the Project Baseline Health Study

**DOI:** 10.1101/2024.12.20.24319449

**Authors:** Jessica A. Regan, Jordan Franklin, Kalyani Kottilil, Nicholas Cauwenberghs, Kenneth W. Mahaffey, Pamela S. Douglas, Fatima Rodriguez, Francois Haddad, Adrian F. Hernandez, Svati H. Shah, Lydia Coulter Kwee, the Project Baseline Health Study Group

## Abstract

The heart is a metabolically active organ with high energy demands that depend on mitochondrial oxidative phosphorylation, however as the heart begins to fail there is metabolic inflexibility and changes in mitochondrial function. Epigenetic changes such as DNA methylation can modify the transcription of genes essential for normal mitochondrial function, however this has not been studied in Stage B or pre-heart failure (HF). In 752 participants from the Project Baseline Health Study, we tested differences in DNA methylation in mitochondrial-related genes for association with Stage B HF compared to Stage A (at-risk for HF). We then validated these findings in 918 participants from the CATHGEN study for risk of incident HF hospitalization. One region on chromosome 13 overlapping with the cysteinyl-tRNA synthetase 2 (*CARS2*) gene, containing five cytosine-phosphate-guanine (CpG) probes, was significantly associated with a small mean regional increase in DNA methylation in Stage B HF (0.63%, Stouffer p=0.005). In CATHGEN, one *CARS2* probe, cg08289839, was significantly associated with time-to-incident HF hospitalization (adjusted HR 1.74 [95% CI 1.18-2.56], FDR p=0.02). In this study of epigenetic changes of mitochondrial genes, these results indicate that *CARS2* DNAm may play a role in HF pathogenesis and should be studied in future HF research.

---

A 2021 consensus statement defined four stages of heart failure (HF): at-risk for HF (Stage A), pre-HF (Stage B), HF (Stage C) and advanced HF (Stage D), however, there is limited understanding of mechanisms by which individuals transition from Stage A to Stage B and to then symptomatic HF^1^. The heart’s high energy demands are dependent on mitochondrial oxidative phosphorylation of fatty acids for the majority of ATP generation, while HF is characterized by metabolic inflexibility and increased reliance on alternative fuel substrates. Epigenetic changes such as DNA methylation (DNAm) can modify transcription of genes central to mitochondrial function, thereby impacting downstream cellular and metabolic function. DNAm changes have been identified in myocardial tissues of subjects with end-stage HF, but whether mitochondrial-related DNAm may signal early molecular changes in Stage B HF has not been studied^2^. Here, we investigate the role of DNAm in mitochondrial-related genes in early-stage HF and in risk of incident HF hospitalization.

The Project Baseline Health Study (PBHS) recruited 2502 participants, of which 752 had either Stage A or Stage B HF along with peripheral blood DNAm profiling from the Illumina EPIC array^3^. Mean age was 54.4 ±15.8 years, 46.9% of participants were male and 39.9% self-identified as non-white race. 509 participants were at-risk for HF (Stage A) defined as history of hypertension, prevalent cardiovascular disease, diabetes, obesity, chronic kidney disease, prior exposure to cardiotoxins or family history of cardiomyopathy without structural heart disease. 243 had Stage B HF, defined as subjects without current or prior signs or symptoms of HF, but with at least one of the stage A HF characteristics, along with evidence of structural heart disease including left ventricular mass, hypertrophy, chamber enlargement, valvular heart disease or systolic or diastolic cardiac dysfunction on echocardiography^4^. We considered 25,774 cytosine-phosphate-guanine (CpG) probes in 1,192 mitochondrial-related genes identified from MitoCarta 3.0, Harmonizome 3.0, the PANTHER database and genes previously described to have DNAm changes or variable expression in HF^2^. Linear regression was used to test the association of Stage B (vs. stage A) HF with DNAm using the *limma* package for individual probes and *DMRcate* for region-based analysis (R v4.4.2). Models were adjusted for age, self-reported race, sex, body mass index, systolic blood pressure, creatinine, hypertension, diabetes, coronary disease and cell proportions derived from a complete blood count. Significance was considered at a false discovery rate (FDR) p<0.05 for both individual probe and region-based analyses. Given that hypermethylation is a hallmark of cancer, sensitivity analyses were also performed excluding the 104 participants with a history of cancer. We then tested the association of significant findings with echocardiographic criteria defining Stage B HF. We validated significant findings in the CATHGEN cohort. Although CATHGEN does not have stage B HF phenotyping available, we tested for association between DNAm and time-to-incident HF hospitalization using Cox proportional hazard models in 918 participants without prevalent HF, adjusted for the above covariates. Informed consent was obtained from all participants enrolled in PBHS in accordance with the Helsinki declaration. The study was approved by a central Institutional Review Board (the WCG IRB; approval tracking number 20170163, work order number 1-1506365-1) and IRBs at each of the participating institutions (Stanford University, Duke University, and the California Health and Longevity Institute).

Baseline characteristics of the participants included in these analyses are shown in **Table 1**. No individual mitochondrial-related CpGs were associated with Stage B HF in PBHS after FDR adjustment (**Supplemental Table 1**). In region-based analysis, a 706bp region on chromosome 13 overlapping with the cysteinyl-tRNA synthetase 2 (*CARS2*) gene, which contains five CpGs, was significantly associated with Stage B HF (Stouffer p=0.005), although the mean regional increase in DNAm was small (0.63%) (**Supplemental Table 2**). In sensitivity analyses, excluding participants with history of cancer strengthened the association between the *CARS2* region and Stage B HF (p=2.1×10^−7^) (**Supplemental Table 3, Supplemental Table 4**). There were no significant associations between the five *CARS2* CpG probes and the individual structural and functional components of Stage B HF (**Supplemental Table 5, Supplemental Table 6**). In CATHGEN, baseline characteristics are shown in **Supplemental Table 7**. 254/918 CATHGEN participants had an incident HF hospitalization event. One of the probes in the *CARS2* region, cg08289839, was significantly associated with time-to-incident HF hospitalization: adjusted HR 1.74 (95% CI 1.18-2.56), FDR p=0.02 (**Supplemental Table 8**).

**Table 1.** Baseline Participant Characteristics.

|  |  |
| --- | --- |
| n | 752 |
| Age (mean [SD], years) | 54.4 (15.8) |
| Sex, N male (%) | 353 (46.9) |
| Self-reported Race, N (%) |  |
| American Indian or Alaska Native | 10 (1.3) |
| Asian | 51 (6.8) |
| Black or African American | 184 (24.5) |
| Native Hawaiian or Other Pacific Islander | 7 (0.9) |
| Other | 48 (6.4) |
| White | 452 (60.1) |
| Hispanic, N (%) |  |
| Yes | 66 (8.8) |
| No | 685 (91.1) |
| Don't know | 1 (0.1) |
| Systolic Blood Pressure (mean [SD], mmHg) | 129.6 (15.5) |
| Diastolic Blood Pressure (mean [SD], mmHg) | 79.4 (10.5) |
| Body Mass Index (mean [SD], kg/m<sup>2</sup>) | 31.3 (6.5) |
| Creatinine (mean [SD], mg/dL) | 0.9 (0.3) |
| Coronary Artery Disease, N (%) | 47 (6.2) |
| Hypertension, N (%) | 443 (58.9) |
| Diabetes, N (%) | 171 (22.7) |
| History of Cancer, N (%) | 104 (13.8) |
| Stage B HF, N (%) | 243 (32.3) |

Here, in a study of epigenetic changes of mitochondrial genes, we have shown that *CARS2* DNAm may play a role in HF pathogenesis, as suggested by its association with Stage B HF and HF incidence (**Figure 1**). *CARS2* encodes an aminoacyl-tRNA synthetase that is transcribed by the nuclear genome but exclusively targets mitochondria, where it is imported and enzymatically catalyzes the ligation of cysteine to tRNA molecules. *CARS2* is ubiquitously expressed in the heart and many other tissues, and variants in *CARS2* have been implicated in mitochondrial epileptic encephalopathy, whereas variants in other aminoacyl-tRNA synthetases and nuclear encoded proteins impacting mitochondrial translation have been associated with cardiomyopathy^5^. Here we have used peripheral blood DNAm as a circulating surrogate for myocardial changes in Stage B HF, as unfortunately, access to myocardial tissue in early HF is likely not feasible. Further studies should evaluate the role of epigenetic changes in genes essential to mitochondrial and metabolic function across the spectrum of stages of HF in large scale human cohorts.

**Figure 1.**
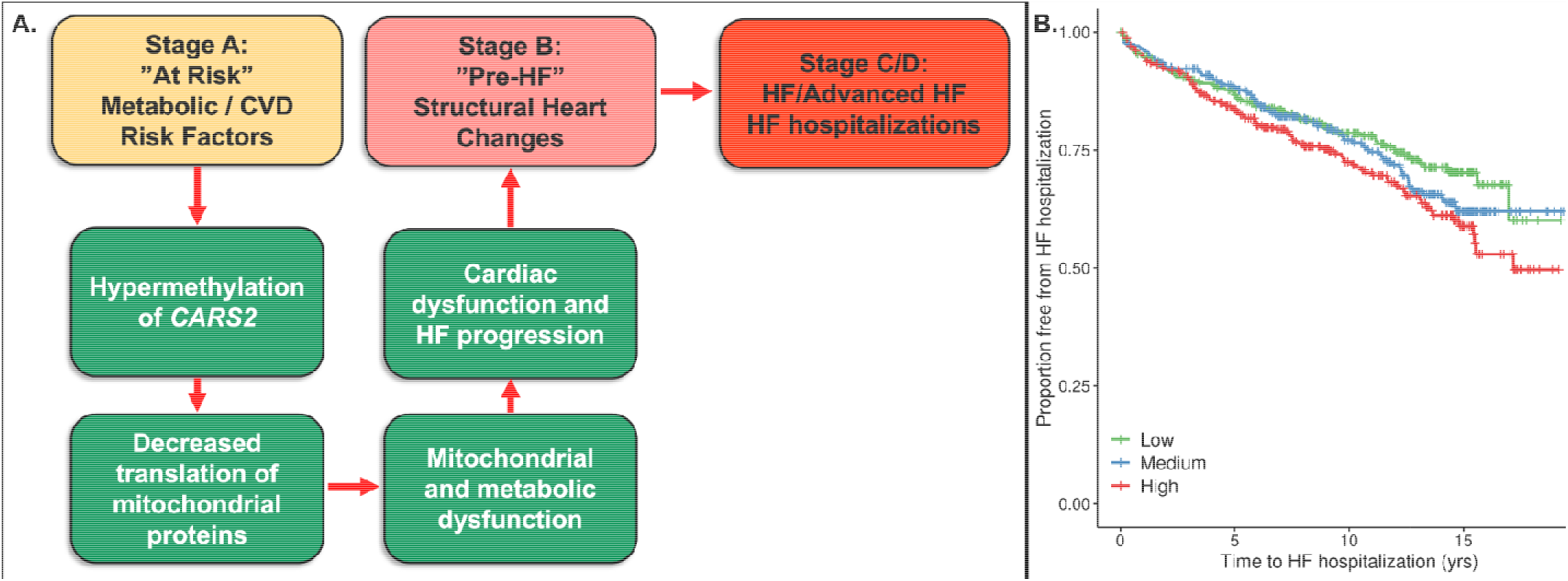
*CARS2* Hypermethylation in Stage B and Incident HF hospitalization. A. Conceptual model exploring the epigenetic changes that may prompt the transition from Stage A HF characterized by cardiovascular and metabolic risk factors to Stage B HF. Mitochondrial promoter hypermethylation of *CARS2* which serves an essential tRNA-synthetase role for mitochondrial protein translation could lead to subsequent metabolic and mitochondrial dysfunction and ultimately subclinical cardiac dysfunction and progressive HF. B. Kaplan-Meier curve by tertile of cg08289839 methylation in *CARS2* in CATHGEN cohort for time-to-HF hospitalization. HF, heart failure; *CARS2*, cysteinyl-tRNA synthetase 2.

## Data Availability

The deidentified PBHS data corresponding to this study are available upon request for the purpose of examining its reproducibility. Requests are subject to approval by PBHS governance.

## ACKNOWLEDGMENTS

We would like to thank the Project Baseline Health Study and CATHGEN participants whose data contributed to this work.

## Ethics Statement

The study was approved by the Duke University and Stanford University Institutional Review Boards. Informed consent was obtained from all participants enrolled in the Project Baseline Health Study in accordance with the Declaration of Helsinki.

## ClinicalTrials.gov Identifier

NCT03154346, https://clinicaltrials.gov/ct2/show/NCT03154346

